# Constructing Parkinson’s disease progression-endotype molecular networks for drug repurposing: multi-omics evidence aggregation and large-scale real-world data validation

**DOI:** 10.1101/2024.01.29.24301961

**Authors:** Manqi Zhou, Alison Ke, Zhenxing Xu, Zhengkang Fan, Kun Chen, Jie Xu, Roberta Marongiu, Fei Wang, Chang Su

## Abstract

Parkinson’s disease (PD) presents with considerable clinical heterogeneity, spanning motor and non-motor symptoms with variable progression trajectories. To investigate the molecular drivers of this variability and identify therapeutic opportunities, we conducted a multi-omics, network-based analysis of the Parkinson’s Progression Markers Initiative (PPMI) cohort, with independent validation in the Parkinson’s Disease Biomarkers Program (PDBP) cohort. By integrating genetic and longitudinal transcriptomic data, we constructed progression-endotype networks, each capturing trait-specific molecular signatures. These networks showed significantly greater connectivity than expected by chance and converged on established PD-associated genes, including Glucosylceramidase Beta 1 (*GBA1*), Apolipoprotein E (*APOE)* haplotype, and Leucine Rich Repeat Kinase 2 (*LRRK2)*. Using these phenotype-informed modules, we applied a network proximity approach to systematically assess 1,595 FDA-approved drugs for repurposing potential. We prioritized 25 candidates, including Zolpidem, Alprazolam, Duloxetine and Primidone. Analysis of real-world clinical data from two large research networks further revealed consistent associations between use of these drugs and reduced incidence of PD-related outcomes. Together, these findings demonstrate the utility of progression-endotype networks for capturing PD progression biology and guiding drug repurposing. This integrative framework connects molecular mechanisms with clinical impact and may inform precision therapeutic strategies for this neurodegenerative disease.

## Introduction

Parkinson’ disease (PD) is the second most prevalent age-associated, progressive neurodegenerative disorder, affecting approximately 2–3% of individuals aged 65 and older^1,2^. PD pathogenesis is still unclear, and the disease remains incurable, with no disease-modifying therapies capable of preventing or reversing its progression^1,3–6^. Clinically, PD presents as a highly heterogeneous disorder, encompassing a broad spectrum of symptoms, including motor impairments such as tremor, rigidity, postural instability, freezing of gait, and speech and swallowing dysfunctions, as well as non-motor problems such as cognitive dysfunction, mood disturbances, sleep disorders, pain, fatigue, and autonomic failures^1,3,4^. Growing evidence emphasizes the considerable inter-individual variability not only in symptom onset but also in the rate and pattern of progression across these distinct clinical dimensions, resulting in highly divergent disease trajectories over time^3,7,8^. Such complexity underscores the urgent need for a comprehensive, multidimensional understanding of PD progression across its full clinical spectrum.

The recent advent of multi-omics offers new opportunities to dissect the molecular underpinnings that govern the heterogeneous progression procedures of PD, with the potential to inform novel therapeutic treatment development ^7^.or instance, a recent genome-wide association study (GWAS) identified the Apolipoprotein (*APOE)* ε4 tagging variant (rs429358, rs7412) as a genetic contributor to cognitive decline and overall PD progression, while gene *ATP8B2* was implicated in motor deterioration^9^. Additional studies have shown that PD patients carrying the Leucine Rich Repeat Kinase 2 (*LRRK2)* G2019S mutation exhibit slower motor progression^10^, whereas those harboring dual mutations in *LRRK2* and Glucosylceramidase Beta 1 (*GBA1*) display accelerated cognitive decline^11^. Moreover, whole blood transcriptomics analyses^12,13^ have uncovered gene signatures linked to PD progression. Despite these advances, prior efforts have mostly focused on individual symptom dimensions, such as motor^9,10,12,13^ or cognitive decline^9,11^, or relied on the composite score derived from a limited set of clinical scales (motor and cognitive assessments)^9^. Importantly, none of these identified susceptible genes for PD progression, along with previously known genes of PD origin^14^, have yet translated into successful development of new PD therapies. This likely reflects the limitations of single-target strategies in tackling a multifactorial disease like PD and highlights the need for therapies that engage multiple disease-relevant pathways or targets.

Emerging network medicine approaches, which integrate multi-omics with the human protein interactome and drug-target networks, have paved the way on translating omics observations into systematical identification of cellular systems and molecular interactome perturbations underling complex diseases, and thereby facilitating in silico drug repurposing^15–19^. Applied in the context of PD, these network-based methodologies offer the potential to comprehensively investigate disease progression across the full spectrum of symptom dimensions, enabling a holistic characterization of the molecular architecture driving divergent disease trajectories, which may thereby advance the prioritization of repurposable drug candidates with the potential to modify PD progression. Meanwhile, the continuously expanding availability of patient-level real-world data (RWD), particularly electronic health records (EHRs), has created unprecedented opportunities to evaluate treatment effects at population scale in real-world clinical settings^20^. RWD encode detailed longitudinal information on treatment exposures and clinical responses, offering insights that may be more directly translatable to patient outcomes^20,21^. The target trial emulation framework, which simulates randomized controlled trials using observational RWD, has emerged as a robust methodological approach for estimating causal treatment effects while rigorously accounting for confounding factors^22^. Integrating RWD-based trial emulation into drug repurposing pipelines enables generation of real-world evidence to validate in silico–predicted candidates^20^, thereby accelerating the translation of molecular discoveries into clinically actionable therapies for PD.

In this study, we developed an integrated pipeline that combines multi-omics, network medicine, and large-scale RWD to pinpoint molecular determinants of PD progression and prioritize repurposing therapeutic candidates aimed at modifying the disease course (Figure 1). We hypothesized that distinct progression-endotypes exist within PD, driving disease progression across different symptom dimensions, and that characterizing these progression-endotypes could inform the development of targeted therapies to prevent or slow PD progression. Specifically, through the analysis of individual-level multi-omics and longitudinal clinical records using our well-established network medicine framework, we systematically constructed progression-endotype modules linked to distinct clinical trajectories in PD. Our findings revealed partially overlapping (e.g. PINK1, *RIT2*) yet distinct gene signatures (*GBA, LRRK2, PRKN, SNCA*, and *FYN*) across endotypes, including both motor and non-motor domains, suggesting that these endotypes reflect divergent molecular mechanisms underlying symptom-specific disease progression. Based on the progression-endotype modules, system pharmacology and network proximity-based approach led to the discovery of Zolpidem, Alprazolam, Duloxetine and Primidone as potential repurposing candidates. The drugs were associated with a reduced incidence of advanced PD outcomes in two large-scale RWD repositories, supporting their potential real-world effectiveness.

**Figure 1.**
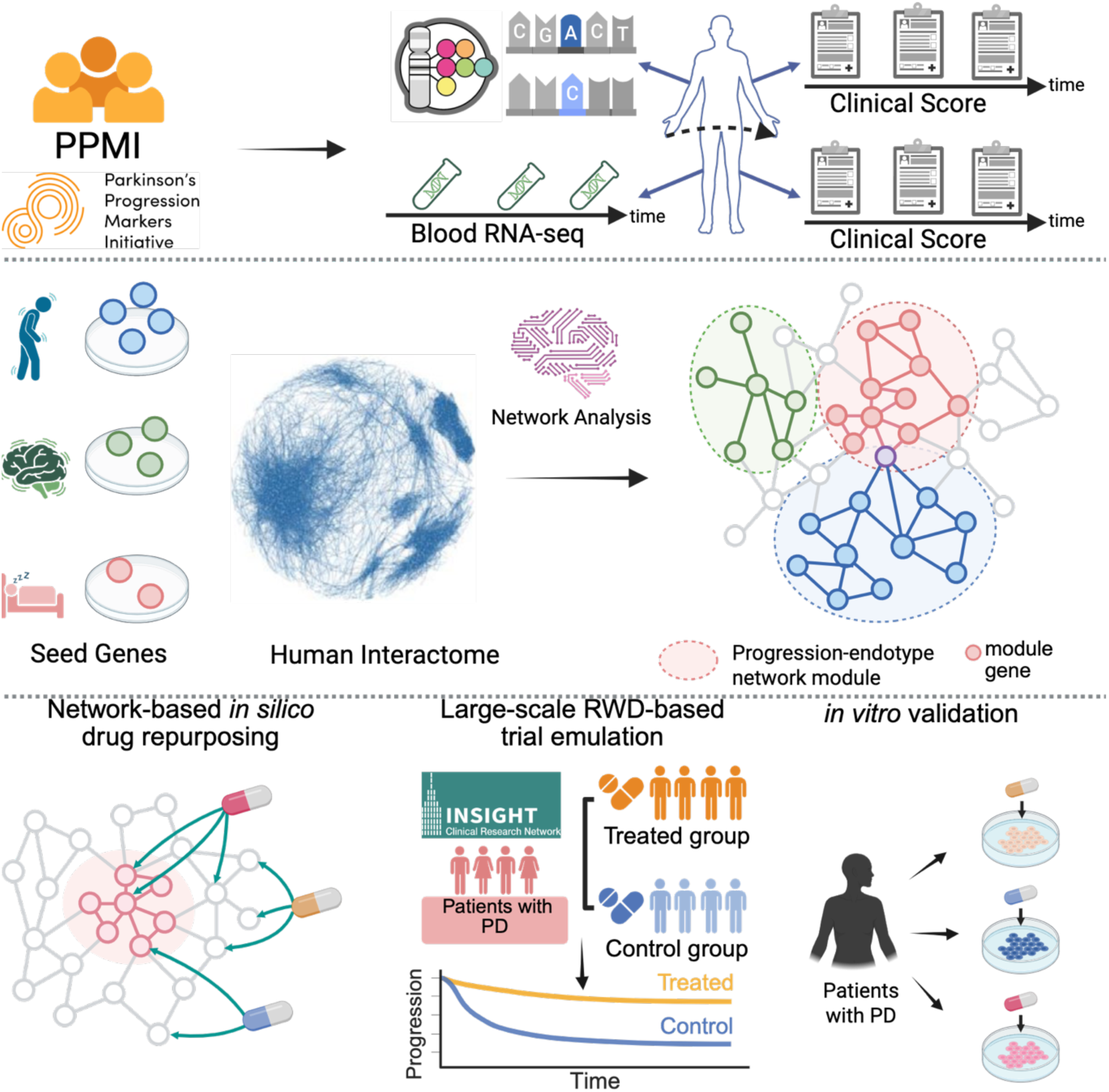
Study pipeline.

## Method

### Study cohorts

We used data from two abundant PD research programs:

1. The Parkinson’s Progression Markers Initiative (PPMI, http://www.ppmi-info.org)^23^, launched in 2010 and sponsored by the Michael J. Fox Foundation, is an international and multi-center observational study dedicated to identifying biomarkers indicative of PD progression. The PPMI study protocol was approved by the institutional review board of the University of Rochester (NY, USA), as well as from each PPMI participating site. Data used in the preparation of this article were obtained on November 2023 from the PPMI database (www.ppmi-info.org/access-data-specimens/download-data), RRID:SCR_006431. For up-to-date information on the study, visit www.ppmi-info.org. This analysis used clinical records, whole exome sequencing data, and whole blood RNA-seq data, obtained from PPMI upon request after approval by the PPMI Data Access Committee. From PPMI, this investigation included participants with *de novo* PD (diagnosed with PD within the last 3 years and untreated at enrollment) for analysis. Participants with less than 2 follow-up visits or 6 months duration after baseline were excluded due to lack of PD progression information.
2. The Parkinson Disease Biomarkers Program (PDBP, https://pdbp.ninds.nih.gov)^24^, established in 2012 and funded by the National Institute of Neurological Disorders and Stroke (NINDS), is an observational study for advancing comprehensive PD biomarker research. More information about the PDBP participants has been described elsewhere^24^. The study protocol for each PDBP site was approved by institutional review board of each participating site. Data of the PDBP cohort were obtained on September 2023 via the Accelerating Medicines Partnership Parkinson’s disease (AMP-PD) Knowledge Platform (http://amp-pd.org) under AMP-PD Data Use Agreement. From PDBP, this study included the participants diagnosed with PD. Similar to PPMI, participants with less than 2 follow-up visits or 6 months duration after baseline were excluded due to lack of PD progression information.

We used PPMI as the development cohort while used PDBP as the independent validation cohort.

### Clinical variables

To study PD progression, we used the following clinical variables:

1. Demographics including age at PD diagnosis onset, sex, race, family history, symptom duration at baseline, and education level.
2. Clinical assessments collected at baseline and follow-up visits:

- Motor manifestations measured by International Parkinson’s disease and Movement Disorder Society-Unified Parkinson’s Disease Rating Scale (MDS-UPDRS)-Part II, MDS-UPDRS-Part III (no treatment or off treatment), total tremor score, postural instability–gait difficulty (PIGD) score^25^, Schwab-England Activities of daily living score.
- Cognition manifestation scales including Montreal Cognitive Assessment (MoCA)^26^, Benton Judgment of Line Orientation (Benton)^27^, Symbol Digit Modalities Test (symbol digit)^28^, Hopkins Verbal Learning Test (HVLT)^29^ for total recall, delayed recall, retention and recognition-discrimination, and semantic verbal-language fluency test (semantic fluency)^30^ and letter-number sequencing (LNS).
- Mood disorders measured by Geriatric Depression Scale (GDS) score^31^ and State-Trait Anxiety Inventory (STAI) score^32^.
- Sleep problems measured by REM Sleep Behavior Questionnaire (RBD)^33^, Epworth Sleepiness Score (ESS)^34^.
- Autonomic dysfunction measured by Scales for Outcomes in Parkinson’s Disease-Autonomic questionnaire (SCOPA)^35^.
3. Cerebrospinal fluid (CSF) biomarkers including phosphorylated tau, total tau, Aβ_42_, α-synuclein collected at baseline and follow-up visits.
4. PD medicine: we used levodopa equivalent daily dose (LEDD) at visit.

For the PPMI cohort, we used all the variables listed above. For the PDBP cohort, we included only the motor assessments, MoCA, and ESS due to missing data or a missing rate exceeding 50%.

### Clinical data processing

We calculated total score for each clinical assessment as mentioned above. The MoCA scores underwent inverse scoring to represent the count of incorrect responses out of a total possible score of 30. Similarly, semantic fluency was inversely scored against the highest individual score recorded at baseline. Each clinical score was z-score normalized based on its mean and standard deviation at the baseline visit.

### Genetic data analysis

We used whole genome sequencing (WGS) data of participants. Of note, identifying statistically and biologically significant signals from the whole genome is challenging due to the small effect sizes of single mutations and limited sample size of individuals with well-phenotyped, longitudinal progression information of PD. Therefore, we focused on loci reported in previous genome-wide association studies (GWAS) of PD, PD progression, PD-related pathology (e.g., a-synuclein measurement), and other neurodegenerative disorders (including Alzheimer’s disease, Huntington’s disease, dementia, and Lewy body dementia), collected from the GWAS Catalog^36^. GWAS studies that focused on “Young adult-onset Parkinsonism”, “age at onset, Parkinson disease”, “pesticide exposure measurement, Parkinson disease”, “age at diagnosis, Parkinson disease”, “Lewy body measurement, Parkinson disease”, “motor function measurement”, “drug-Induced dyskinesia, response to levodopa”, “impulse control disorder”, “mortality”, were excluded for analysis. As such, we obtained a total of 2,972 risk loci.

We then performed genetic analysis with linear mixed-effects models to identify genetic loci that are associated with PD progression speed. More specifically, for each tested SNP, we used the SNP×time term to examine its association with PD progression speed measured by a specific clinical score. Individuals were considered as random effects. In addition, we included covariates including age at PD onset, race, symptom duration, LEDD at visit, LEDD× *t* interaction, and value of the tested clinical score at the baseline visit; we also included education years as an additional covariate when measuring cognitive progression. As such, we calculated *P* value and *β* of the SNP×time term. Variants with *P* value < 0.05 were mapped to genes using the “Mapped gene(s)” field from the GWAS Catalog, which includes genes overlapping the variant or the nearest 5’ and 3’ genes if intergenic. These mapped genes were considered as seed genes for PD progression and fed to the network medicine algorithm for gene module identification, which were detailed below.

### Transcriptomic data analysis

We used whole-blood bulk RNA-seq data of participants. Genes were annotated using UniProt^37^, and only protein coding genes were included for analysis. Genes with low expression levels across all samples were excluded. We kept genes with more than 100 counts in at least 500 visits. To identify genes whose expression evolving patterns are associated with progression of a specific clinical score, we used the following two-stage procedures:

First, we applied a linear mixed-effects model to estimate individual-level progression rates in each specific clinical score. Here, the time term was considered as the explanatory variable of interest. Individuals were considered as random effects. We included covariates as mentioned above. After fitting, the model estimated a progression rate for each individual in the specific clinical score.

Second, similarly, we applied linear mixed-effects model to estimate individual-level progression rates in expression level of each specific gene.

Third, we examine association between the progression rate of each clinical score and that of each gene, using a generalized linear model (GLM). Genes with *P* values < 0.05 were selected as seed genes for PD progression and fed to the network medicine algorithm for gene module identification, which were detailed below.

### Construction of human protein-protein interactome network

We assembled commonly used human protein-protein interactome (PPI) databases with experimental evidence and in-house systematic human PPIs to build a comprehensive human PPI network. PPI databases we used included: (i) kinase-substrate interactions via literature-derived low-throughput and high-throughput experiments from Human Protein Resource Database (HPRD)^38^, Phospho.ELM^39^, KinomeNetworkX^40^, PhosphoNetworks^41^, PhosphositePlus^42^, and dbPTM 3.0^43^; (ii) binary PPIs from 3D protein structures from Instruct^44^; (iii) binary PPIs assessed by high-throughput yeast-two-hybrid (Y2H) experiments^45^; (iv) protein complex data (∼56,000 candidate interactions) identified by a robust affinity purification-mass spectrometry collected from BioPlex V2.0^46^; signaling networks by literature-derived low-throughput experiments from the SignaLink2.0^47^; and (vi) literature-curated PPIs identified by affinity purification followed by mass spectrometry from BioGRID^48^, HPRD^49^, InnateDB^50^, IntAct^51^, MINT^52^, and PINA^53^. In total, 351,444 PPIs connecting 17,706 unique proteins are now freely available at the AlzGPS website (https://alzgps.lerner.ccf.org/)^54^. In this study, we utilized the largest connected component of this dataset, including 17,456 proteins and 336,549 PPIs. The PPI network was used for network analyses of genetic and transcriptomic data for PD progression-endotype molecular module identification, which were detailed below.

### Network-based approach for gene module identification

We next conducted network analyses that leveraged genetic and transcriptomic findings associated with PD progression to derive PD progression-endotype gene modules. Here, we posit that a PD progression-endotype gene module are the potential drivers of PD progression in a specific symptom dimension. To achieve this, we used our Genome-wide Positioning Systems network (GPSnet) algorithm^17,19,55^, which demands two inputs:

1. The PPI network built, which were detailed above.
2. Seed genes along with their risk scores obtained from omics signals. Specifically, i) for genetic signals, we first mapped seed variants associated with PD progression to their corresponding gene symbols based on GWAS summary statistics obtained from the GWAS Catalog. As such, a list of seed genes was generated for each clinical score. The risk score *s*(*i*) for each gene *i* was defined as the maximum value of |-log_10_(*P* value) **×** *β|* across all mapped variants. ii) For transcriptomic signals, we used the seed genes derived using RNA-seq data. The risk score *s*(*i*) for each gene *i* was defined as |-log_10_(*P* value) **×** *β|*. For all non-seed genes, we defined *s*(*i*) = 0.

The GPSnet algorithm then generated gene modules using the following procedures. First, a random walk with restart (RWR) procedure was implemented to obtain the smoothed risk score ŝ(*i*) for each gene *i* within the whole PPI network. Next, the following procedures was adapted to construct a gene module *M*: It first initialized module *M* only containing a randomly selected seed gene and then expanded the module by involving genes one by one, while enhancing gene connectivity within the module (Criterion I) and improving module level gene score (Criterion II). Specifically, it included a gene *i* ∈ Γ_*M*_ into *M* (Γ_*M*_ is a set of genes linking to any genes within current module *M*) if *P*(*i*) ≤ 0.01, calculated with Eq. (1), indicating genes within *M* densely connect to each other after including *i* (Criterion I) and *S*(*M* ∪ {*i*}) > *S*(*M*), calculated with Eq. (2), indicating module-level risk score increased after involving gene *i* (Criterion II).

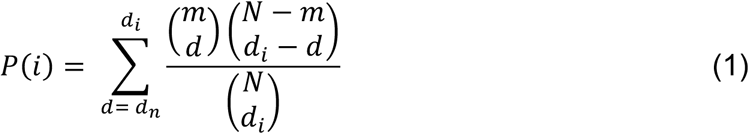

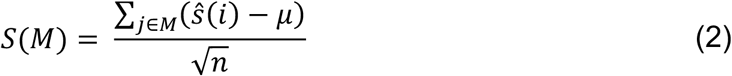

where, *m* denotes the number of genes in module *M*, *N* is the total number of genes in the PPI, *d_n_* is the number of neighbors of gene *i* within the module *M*, *d_i_* is the degree of gene *i*; μ denotes the average risk score of all genes in the PPI network, and *S*(*M*) denotes the module-level gene risk score of the module *M*.

We stopped module expansion when *S*(*M*) cannot be increased or *P*(*i*) ≥ 0.01 by involving new genes. In this study, we repeated above procedures multiple 100,000 times and obtained a collection of raw modules. We generated the final gene module by assembling the raw modules with the top 0.5% highest Γ_*M*_ scores and define the largest connected component after discarding isolated genes.

In this study, we ran GPSnet algorithm based on genetic and transcriptomic findings separately to obtain gene modules of a specific progression-endotype, e.g., cognitive progression measured by MoCA. We then combined genetic- and transcriptomic-based modules to construct the progression-endotype module.

### Pathway enrichment analysis

Pathway enrichment analyses were performed with g:Profiler^56^ using Gene Ontology (GO)^57^, Kyoto Encyclopedia of Genes and Genomes (KEGG)^58^, and Reactome^59^ databases. From the GO database, we used biological process terms. Multiple correction was conducted by controlling false discovery rate (FDR) and the FDR adjusted *P* values < 0.05 were considered as significant. The significant GO biological process terms enriched in each progression-endotype gene module were further refined using GOxploreR ^39^ and REVIGO^40^, eliminating GO terms with redundant biological information. Last, we manually clustered and annotated the final GO terms for better interpretation.

### Construction of drug-target network

We assembled physical drug-target interactions for United States Food and Drug Administration (FDA)-approved drugs via integrating six commonly used data sources^17,19,55^. These data sources include DrugBank^60^, BindingDB^61^, ChEMBL^62^, Therapeutic Target Database^63^, PharmGKB^64^ and IUPHAR/BPS Guide to PHARMACOLOGY^65^. A physical drug-target interaction was defined based on binding affinities K_i_ (inhibition constant/potency), K_d_ (dissociation constant), IC_50_ (median inhibitory concentration), or EC_50_ (median effective concentration) ≤ 10 μM. We retained only those items satisfying the following criteria: (1) K_i_, K_d_, IC_50_ or EC_50_ ≤10 μM; (2) protein targets having a unique UniProt accession number; (3) protein targets marked as ‘reviewed’ in the UniProt database; and (4) proteins present in Homo sapiens. In total, we obtained 15,367 drug-target interactions connecting 1,608 FDA-approved drugs and 2,251 unique human targets/proteins.

### Network proximity calculation for drug repurposing

Following previous studies^17,19,66^, we defined the average shortest path distance *d*;(*S*, *T*) between a gene module *S* (e.g., a PD progression-endotype module) and the set of targets *T* of a tested drug, as follows:

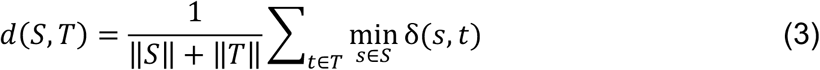

where, ‖*S*‖ and ‖*T*‖ denote size of *S* and *T*, respectively; δ(*a, b*) indicates the shortest path between gene/protein *a* and target *b* within the PPI network. Next, a permutation test was used to evaluate significance of network-based distance between the gene module and each tested drug candidate. Specifically, we first generated a reference distance distribution corresponding to the expected distance between the tested drug’s target set *T* and a random drawn gene set *S*_-_ with the degree (number of neighbors of a gene in the network) distribution and size of module *S*. We repeated the procedure 1000 times to obtain a set of *S*_-_. A Z-score was calculated by using the mean and standard deviation of the reference distribution to normalize the observed distance between *S* and *T*.

### Randomized trial emulation based on large-scale real-world data (RWD)

We conducted randomized trial emulation using large-scale RWD to evaluate treatment effects of the predicted drug candidates.

#### RWD sources

We used two independent large-scale RWD repositories:

1. INSIGHT Clinical Research Network (CRN)^67^. The INSIGHT CRN, supported by the Patient-Centered Outcomes Research Institute (PCORI), includes longitudinal clinical records for over 15 million patients in the five top academic medical centers in NYC metropolitan area, including Weill Cornell Medicine, Albert Einstein School of Medicine/Montefiore Medical Center, Columbia University Irving Medical Center, Icahn School of Medicine/Mount Sinai Health System, and NYU Grossman School of Medicine/NYU Langone Health. The use of the INSIGHT data was approved by the Institutional Review Board (IRB) of Weill Cornell Medicine under protocol 24-07027736.
2. OneFlorida+ Clinical Research Consortium^68^. OneFlorida+ is another CRN supported by PCORI, which includes 12 healthcare organizations and contains longitudinal and linked patient-level data. This study used de-identified, robust linked patient-level RWD of 17 million patients in Florida, 2.1 million in Georgia (via Emory), and 1.1 million in Alabama (via UAB Medicine) since 2012 and covering a wide range of patient characteristics including demographics, diagnoses, medications, procedures, vital signs, and lab tests. The use of the OneFlorida+ approved by the IRB of University of Florida under protocol IRB202300639.

#### Eligibility criteria

Patient eligibility criteria for analysis included:

1. Patients should have at least one PD diagnosis according to International Classification of Diseases 9th and 10th revision (ICD-9/10) codes, including 332.0 (ICD-9) and G20 (ICD-10).
2. Age > 50 years old at the first PD diagnosis event.
3. Patients who had neurodegenerative disease diagnoses before his/her first PD diagnosis was excluded.

#### Treatment strategies

For each emulated trial, we compared the two treatment strategies: 1) receiving a tested drug vs 2) receiving an alternative drug of the tested drug. Here, we defined an alternative treatment in two ways: i) a randomly selected drug except for the tested drug, or ii) a randomly selected drug from the same ATC-L2 classification with the tested drug except for the tested drug.

#### Outcomes

We measured the outcomes including dementia and falls (indicating advanced motor impairment and dyskinesia)^17,69,70^, which were defined by ICD-9/10 diagnosis codes. Drug treatment efficiency was defined by reducing the risk of onset of the outcomes.

#### Study follow-up

Each patient was followed from his/her baseline until the day of the first outcome event, or loss to follow-up (censoring), whichever happened first.

#### Trial emulation

We retrieved the DrugBank ID for each tested drug and converted it to RxNorm and National Drug Code (NDC) codes. We defined the PD initiation date of each patient as six months prior to his/her first recorded PD diagnosis event^17,70^, accounting for the likelihood that PD may be latently present before formal diagnosis. We then defined *Time Zero* as the start of treatment with a tested drug or its alternative. We required that the *Time Zero* occurred after the PD initiation date and before onset of examined outcomes. We defined the baseline period as the time frame before *Time Zero*.

For each tested drug, we defined the treated group as eligible PD patients who initiated the tested drug after their PD initiation. Our analyses were restricted to drugs that were used by at least 100 PD patients. We then built the control group by selecting PD patients who received an alternative treatment defined above. This study used an intention-to-treat approach, so patients remained in their initially assigned treatment group regardless of the discontinuation of the prescribed treatment^71–73^.

We assumed that the treated group and control group were exchangeable at baseline conditional on baseline covariates. Based on previous studies^17,69,70^ and clinical experience, we included the following baseline covariates: (1) demographics including age, gender, and race, as well as the time from the PD initiation date to the drug index date (i.e., *Time Zero*). (2) 64 comorbidities including comorbidities from Chronic Conditions Data Warehouse and other risk factors that were selected by experts. The comorbidities were defined by a set of ICD-9/10 codes. (3) Medication prescriptions. We grouped drug prescriptions coded as NDC or RXNORM codes into their major active ingredients coded in RxNorm defined in UMLS^74^. We included usage of 200 most prevalent prescribed drug ingredients as covariates in this analysis. In total, 267 covariates were included for analysis.

For each emulated trial, we applied a propensity score method^75^ to address confounding effects. Specifically, we estimated a propensity score for each individual using logistic regression with ridge penalty, based on the baseline covariates detailed above, followed by 1:1 nearest-neighbor matching^76^. The covariate balance after matching was assessed by absolute standardized mean difference (SMD). For each covariate, it was considered balanced between the treated group and control group, if SMD ≤0.2. Further, a trial is balanced if the number of unbalanced covariates is no more than 2% of total covariates.

For each emulated trial, we used a propensity score framework to learn the empirical treatment assignment given the baseline covariates and used an inverse probability of treatment weighting to balance the treated and control groups^76^. A 1:1 nearest-neighbor matching was performed to build the matched control group^76^. The covariate balance after propensity score matching was assessed using the absolute standardized mean difference (SMD)^77^. For each covariate, it was considered balanced if its SMD≤0.2, and the treated and control group were balanced if only no more than 2% covariates were not balanced. To enhance robustness of the analysis, we created 100 emulated trials for each tested drug. Tested drugs that had <10 successfully balanced trials were excluded for analysis. As such, for each tested drug, we emulated: 1) A pool of balanced trials (n=10) adopting random drugs as alternative, 2) A pool (n=10) adopting ATC-L2 drugs as alternative, and 3) A pool combining trials in 1) and 2) (n=20).

#### Treatment effect estimation

For each tested drug, we estimated drug treatment effect for each balanced trial by calculating the hazard ratio (HR) using a Cox proportional hazard model, comparing the risk to develop a specific outcome between the treated and control groups. We reported the median HR with 95% confidence intervals (CI) obtained by bootstrapping^78^. A HR < 1 indicated the tested drug can reduce risk to develop a specific outcome and a P value < 0.05 was considered as statistically significant.

The trial emulation pipeline for treatment effect estimation was implemented using Python packages psmpy^79^ for propensity score framework and lifelines^80^ for the Cox proportional hazard model.

## Result

### Patient data

In this investigation, we used data from two multi-institutional research programs: the Parkinson Progression Marker Initiative (PPMI) study^23^ and Parkinson Disease Biomarkers Program (PDBP)^24^. From the PPMI, we included a total of 500 *de novo* PD participants (diagnosed with PD within the last 3 years and untreated at enrollment), comprising 209 (41.8%) females and 291 (58.2%) males, with an average age of 59.7 ± 9.76 years at PD diagnosis onset. All included participants have baseline records and records collected throughout the 14.23 ± 3.73 visits of 8.72 ± 2.54 years follow-up. To examine molecular underpinnings of PD progression, we used demographics, genetic data, time-course whole blood RNA seq, and longitudinal clinical records (motor and non-motor clinical scales and cerebrospinal fluid [CSF] biomarkers). See Methods and Table S1.

From the PDBP, we included a total of 223 PD participants (diagnosed with PD within the last 3 years and untreated at enrollment), comprising 90 (40.4%) females and 133 (59.6%) males, with an average age of 61.6 ± 9.86 years at PD diagnosis onset. All included participants have baseline records and records collected throughout the 6.31 ± 2.21 visits of 2.85 ± 1.03 years follow-up. To examine molecular underpinnings of PD progression, we used demographics, genetic data, time-course whole blood RNA seq, and longitudinal clinical records (motor and non-motor clinical scales). See Methods and Table S2. The PDBP data were used for independent validation.

Detailed clinical characteristics of participants studied can be found in Table 1.

**Table 1.** PPMI cohort summary statistics.

|  |  |  |  |
| --- | --- | --- | --- |
| # patient |  |  | 500 |
| # visit,<br>mean(SD) |  |  | 14.23(3.73) |
| visit time<br>gap, year,<br>mean(SD) |  |  | 8.72(2.54) |
| Demographic | Age onset, mean(SD) |  | 59.7(9.76) |
|  | Sex male, N(%) |  | 291 (58.2) |
|  | Race white, N(%) |  | 482(96.4) |
|  | Symptom duration, month,<br>mean(SD) |  | 38.77(43.22) |
|  | Family history, N(%) |  | 145(29) |
|  | Education history group, N(%) | <12 years | 48 (9.6) |
|  |  | 12 - 16 years | 260 (52) |
|  |  | >16 years | 192 (38.4) |
| Medicine | Levodopa equivalent daily dose |  | 523.94 |
| Motor | Motor | UPDRSII, mean(SD) | 9.11(6.54) |
|  |  | UPDRSIII, mean(SD) | 24.84(11.85) |
|  |  | Schwab England<br>score, mean(SD) | 87.07(11.68) |
|  | Combined motor | Tremor score, mean<br>(SD) | 0.51(0.41) |
|  |  | PIGD score,<br>mean(SD) | 0.39(0.44) |
| Non-motor | Cognition | MoCA, mean(SD) | 26.51(3.44) |
|  |  | Benton, mean(SD) | 25(4.80) |
|  |  | HVLT, mean(SD) | 47.07(11.46) |
|  |  | LNS, mean(SD) | 11.26(2.97) |
|  |  | Semantic fluency,<br>mean(SD) | 51.5(10.99) |
|  |  | SDT, mean(SD) | 44.83(10.70) |
|  | Mood | GDS, mean(SD) | 2.9(2.98) |
|  |  | STAI, mean(SD) | 66.42(19.04) |
|  | Autonomic | SCOPA, mean(SD) | 19.84(13.13) |
|  | Sleep | REM, mean(SD) | 5.01(3.35) |
|  |  | ESS, mean(SD) | 7.07(4.43) |
| Biomarker |  | p-tau | 14.07(5.21) |
|  |  | t-tau | 166.49(59.25) |
| | | Aβ<sub>42</sub> | 914.45(370.77) |
| | | αSyn | 1472.17(608.31) |

### PD-progression endotype modules

We first investigated genetic factors associated with PD progression rates. Recognizing the small effect size of individual variants and the limited availability of well-phenotype longitudinal data, we focused on 2,972 loci previously implicated in PD and related pathologies curated from the GWAS Catalog (Method, Table S3). Using a linear mixed-effects model over 13 years of follow-up, we identified an average of 157 SNPs per trait that are significantly associated with progression rates (P value < 0.05; Table S4; Figure S1a). To evaluate robustness, we applied the same analytical pipeline to the PDBP cohort, observing broadly consistent SNP-time interactions across datasets (Figure S1b). Identified associated variants were mapped to either overlapping or nearest genes and subsequently used as input for the Genome-wide Positioning Systems network (GPSnet), which identifies trait-specific genetic modules by optimizing gene connectivity and cumulative module-level risk scores (Methods). This network-based approach yielded genetic modules with an average of 180 genes per trait (Table S5, Figure S2a).

Recognizing that genetic variation alone may not fully capture the molecular architecture of disease progression^81^, we next incorporated longitudinal transcriptomic data to complement the genetic findings. We analyzed whole-blood RNA-seq profiles from PD participants using a two-stage modeling strategy (see Methods). In the first stage, we estimated individual-specific progression slopes for each clinical trait using linear mixed-effects models, with time as the primary explanatory variable and individual-level random effects, adjusting for relevant covariates. In the second stage, we applied the same modeling strategy to each protein-coding gene excluding genes with consistently low expression. Associations between clinical and gene-level progression rates were then evaluated using a generalized linear model, with genes showing nominal significance (P < 0.05) retained as trait-associated transcriptomic seed genes. These genes served as input to GPSnet, which identified transcriptomic modules associated with each clinical trait by optimizing gene connectivity and module-level relevance. Similarly, we applied the same analytical pipeline to the PDBP cohort, observing broadly consistent gene-time associations across datasets (Figure S1c). The resulting modules had an average size of 220 genes, ranging from 120 genes for the Benton test to 353 genes for MoCA (Figure S2b, Table S5).

To construct gene modules for PD progression-endotypes, we integrated genetic and transcriptomic modules by taking their union, generating progression-endotype-specific networks that capture a broader spectrum of disease-associated gene interactions (Figure 2, Table S6). Specifically, we combined the UPDRSII, UPDRSIII, and Schwab-England Activities of daily living score into a unified motor endotype module. Among cognitive clinical traits, we prioritized MoCA as the primary outcome and used the other measures as supplementary evidence (Figure 2c). The integration of genetic and transcriptomic signals provides a more comprehensive characterization of the PD progression endotype modules. For the motor endotype (Figure 2a), modules derived from genetic signals exhibit significantly higher node degrees compared to the reference protein–protein interaction (PPI) network (median degree: 96 for the genetic module versus 18 for the PPI; one-sided Wilcoxon test, *P* = 7.78 × 10^-60^, Fig. 3a). A similar pattern is observed in the motor transcriptomic module (median degree: 62.5; one-sided Wilcoxon test, *P* = 1.55 × 10^-56^), indicating that both genetic and transcriptomic modules form tightly connected subnetworks enriched for disease-relevant interactions. Notably, genes shared between the genetic and transcriptomic modules show substantially higher connectivity than those unique to either source (median degree: 417; overlap versus genetic, *P* = 4.53 × 10^-43^; overlap versus genomic, *P* = 1.32 × 10^-50^). A similar trend is observed in the MoCA endotype modules (Figure 2b), where the genetic (median: 198) and transcriptomic (median: 139.5) modules both exceed the PPI background (median: 18), and the overlapping module again displays the highest degree (median: 535; genetic versus PPI, *P* = 3.60 × 10^-63^; transcriptomic versus PPI, *P* = 5.72 × 10^-31^; overlap versus PPI, *P* = 3.03 × 10^-34^). All progression-endotype modules (Figure S3) exhibited significantly higher node degrees than the PPI background (adjusted p value < 0.05), along with significantly greater connectivity in the overlapping modules compared to their respective source modules (adjusted p value < 0.05). These findings suggest that genomics and transcriptomic signals capture distinct yet complementary dimensions of the disease, with their convergence/overlap highlighting core components of the underlying of Parkinson’s disease–associated molecular networks. The identified modules exhibit higher edge density and node closeness than randomly permutated sub-nets from the PPI network, suggesting that these modules form tightly connected subnetworks. The edge density is 0.05 (Z-score 47.16) for the motor module, 0.11 (Z-score 3.03) for the MoCA module, and has adjusted p value < 0.001 comparing with random networks (Figure 2d). Notably, overlapping genes have an even higher edge density (0.07, Z-score 56.94), reinforcing their central role in PD progression (Figure S4, Table S7). These shared genes also exhibit higher network density and lower network closeness, further emphasizing their functional importance in bridging multiple phenotypes.

**Figure 2.**
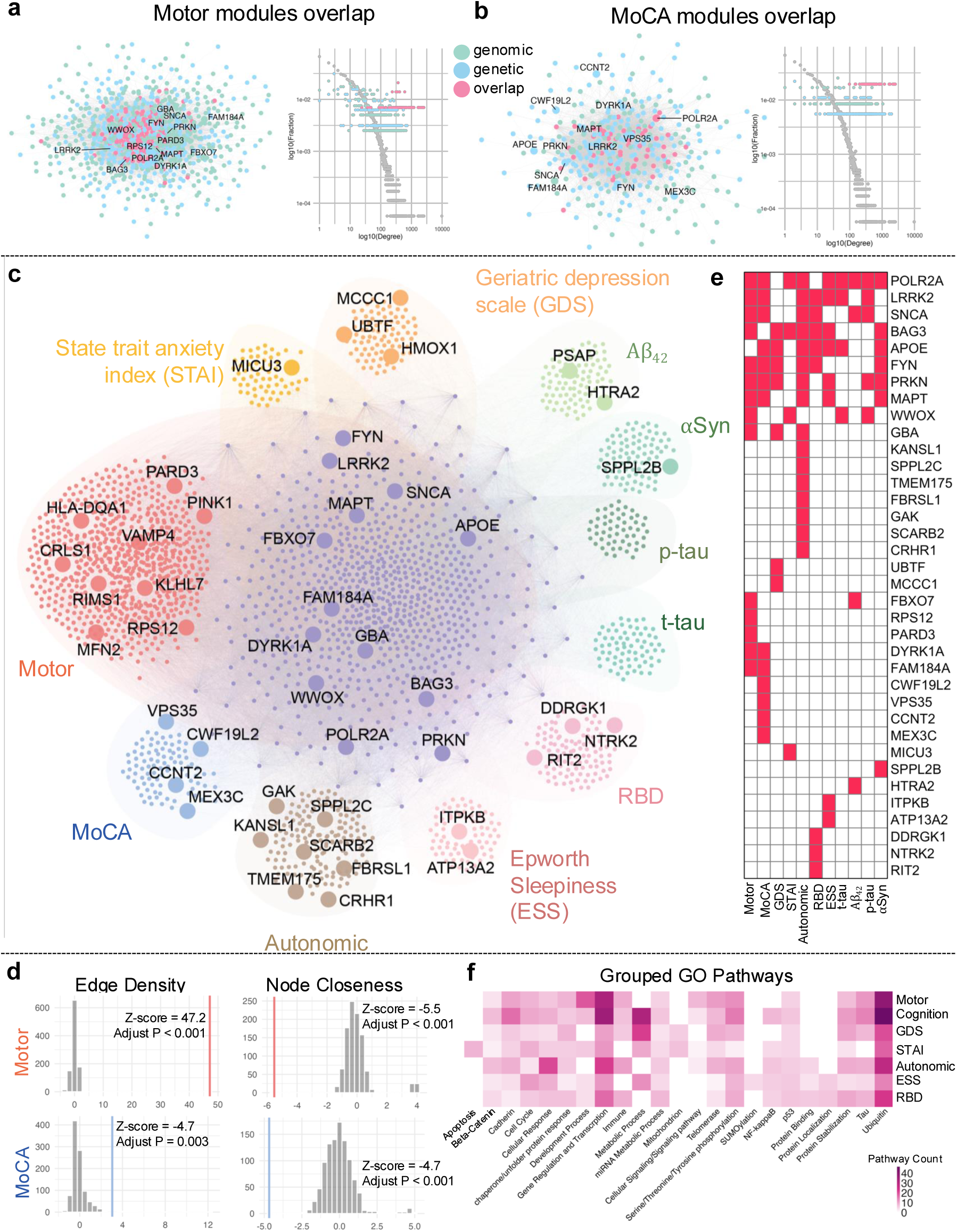
PD progression-endotype modules.

**Figure 3.**
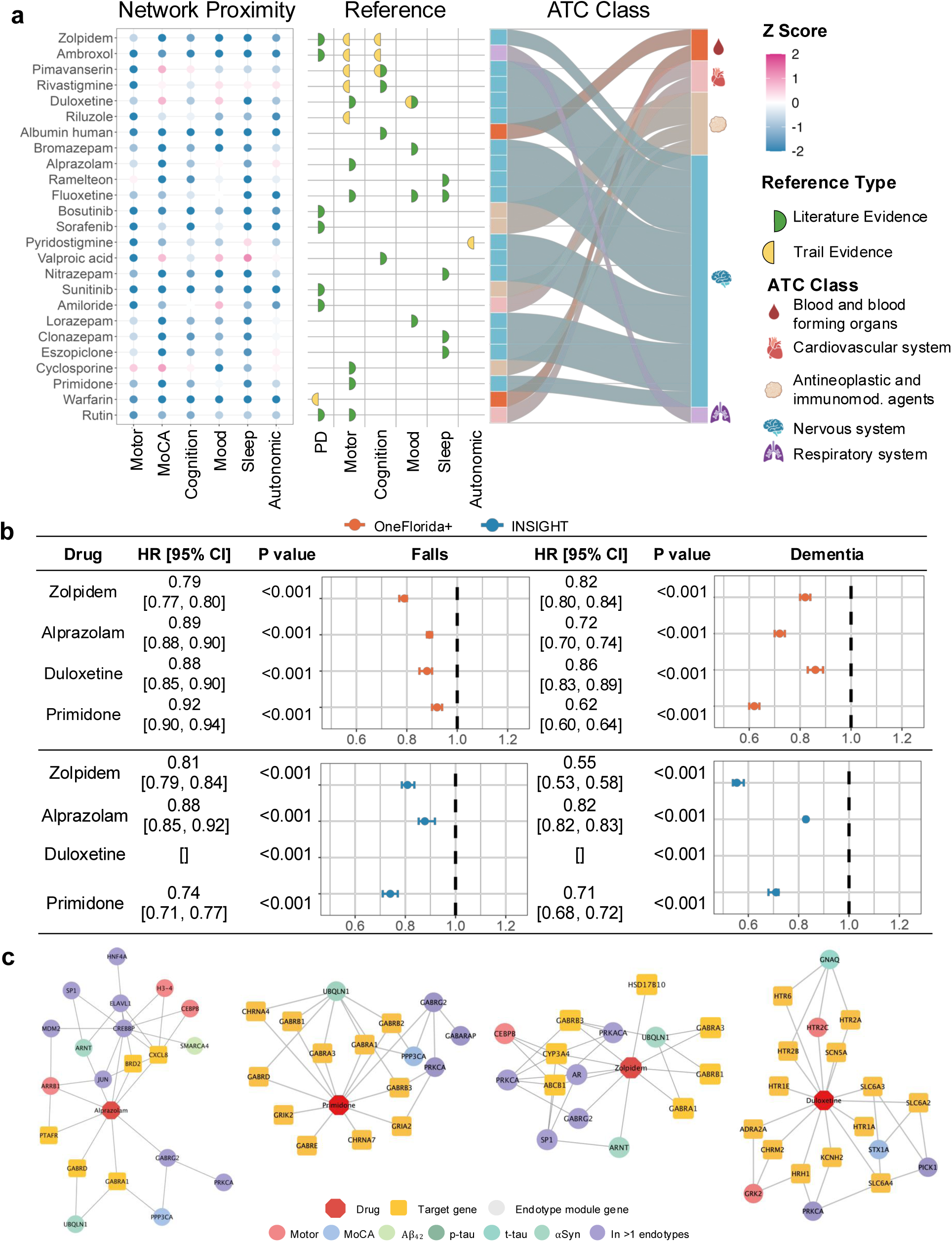
Network-based drug repurposing and real-patient data calidation.

Interestingly, progression endotype-specific genes revealed distinct functional roles across PD progression. In the motor progression-endotype modules (854 genes in total), *PINK1* (associated with Tremor Score endotype) and *MFN2* (associated with UPDRSIII endotype) emerged as specific molecular drivers. *PINK1* regulates *PARKIN* translocation in impaired mitochondria and promotes their removal via mitophagy^82^. Mutations in *PINK1* cause early-onset parkinsonism characterized by asymmetric onset and tremor-dominant motor symptoms^83^, supporting its relevance to PD motor phenotypes. *MFN2* is essential for mitochondrial fusion and the maintenance of mitochondrial dynamics^84^. In dopaminergic neuron–specific knockout mouse models, loss of *MFN2* leads to movement deficits, including reduced locomotor activity and rearing, and age-dependent motor impairments^85^. These deficits precede dopaminergic neuron loss and are associated with fragmentation of mitochondria in the nigrostriatal pathway.

In the cognition progression-endotype modules (2104 genes in total), *TFEB* (associated with Benton and LNS endotypes) was identified as a cognition-specific regulator. *TFEB* encodes a transcription factor that regulates lysosomal biogenesis and autophagy, processes critical for the clearance of protein aggregates^86^. *TREM2* deficiency has been shown to worsen α-Syn induced microglial lysosomal dysfunction and cognitive impairment in Parkinson’s disease, in part by disrupting TFEB nuclear translocation^87^. *HMOX1* (associated with the GDS endotype; 356 genes) was identified as a depression-specific regulator. Overexpression of HO-1 in transgenic mice induces PD-like neurodegeneration with oxidative stress and dopaminergic dysfunction^88^, while elevated plasma HO-1 levels in PD patients have been linked to reduced hippocampal volume, a region involved in mood regulation^89^. *RIT2* (associated with the RBD endotype; 310 genes) encodes a small GTPase implicated in autophagy regulation and is enriched in a subset of substantia nigra neurons that show selective vulnerability in Parkinson’s disease^90^. *PSAP* (associated with the α-synuclein endotype; 280 genes) encodes a neuroprotective lysosomal protein that regulates α-synuclein clearance. Its overexpression decreases α-synuclein levels in neuroblastoma cells, suggesting a potential role in modulating PD-related protein aggregation^91^.

While genes unique to individual progression endotypes reflect endotype-specific molecular signatures, genes shared across multiple endotype may suggest common biological mechanisms underlying PD progression. Among the shared genes identified across multiple endotypes (Figure 2e), *GBA, LRRK2, PRKN, SNCA*, and *FYN* emerged as central regulators. *GBA* encodes glucocerebrosidase, a lysosomal enzyme involved in glycosphingolipid metabolism. Mutations in *GBA* have been associated with earlier age at onset, faster clinical progression, and increased cognitive impairment in PD via α-synuclein accumulation and lysosomal dysfunction and mitochondrial impairment^92^. *LRRK2* encodes a kinase that regulates α-synuclein accumulation and microglial activation. Its dysregulation contributes to neuroinflammation and neuronal loss in PD through kinase-dependent pathways^93^. *SNCA* and *PRKN* are central to the regulation of α-synuclein clearance and dopaminergic neuron integrity and have been implicated in both familial and sporadic Parkinson’s disease^94,95^. *FYN* encodes a tyrosine kinase involved in immune regulation and neuronal signaling. It has been linked to PD-related processes including α-synuclein phosphorylation, oxidative stress, and neuroinflammation^96,97^.

To evaluate the biological relevance of the identified endotype modules, we conducted pathway enrichment analyses^56^ across six clinical modules (motor, cognition, GDS, STAI, REM, ESS, and SCOPA; Methods). On average, 187 Gene Ontology^57^, 546 Reactome^59^, and 145 KEGG^58^ pathways were significantly per endotype (Figure 2f, Table S8), which were subsequently manually grouped into broader biological categories (Table S9). Ubiquitin-related pathways consistently emerged among the top enriched pathways across all endotypes, highlighting their central role in PD progression. This aligns with prior evidence that components of the ubiquitin–proteasome system regulate α-synuclein aggregation, mitophagy, neuroinflammation, and oxidative stress in PD pathogenesis^98^. Pathways involved in gene regulation and transcription were also highly enriched, suggesting widespread dysregulation of transcriptional control in PD. Notably, tau protein binding was enriched in all endotypes except anxiety, consistent with previous findings reporting no significant association between anxiety symptoms and tau levels^99^. In contrast, apoptosis-related pathways were uniquely enriched in the anxiety module. Given that oxidative stress is a known activator of apoptosis and contributes to dopaminergic neuron degeneration in PD^93,100^, this finding may reflect a stress-associated molecular process underlying anxiety symptoms in PD.

### Progression-endotype network-based PD drug repurposing

We employed an in-silico drug repurposing strategy grounded in network-based proximity analysis to identify potential therapeutic candidates for PD progression (Figure 3, Table S10). Specifically, we calculated network proximity scores between progression-endotype modules and an assembled physical drug-target interactions derived for 1595 FDA-approved drugs from six commonly used data sources (see Methods). In this framework, a negative proximity score indicates a closer network distance between a drug and a disease module, suggesting higher therapeutic relevance. Given the clinical relevance of motor and cognitive decline in PD, we focused our analysis on the motor and cognition endotype modules, in addition to four biomarker modules as they reflect more molecular aspect. This screening identified 396 drugs with negative proximity scores on the motor, cognition/MoCA, and four biomarkers’ modules. To refine the initial screen, we integrated proximity scores with supporting evidence from preclinical studies and published literature, ultimately prioritizing 25 candidate drugs that each had at least one additional line of external support. Among these, 16 belonged to the Nervous System category (ATC Class N), consistent with the central role of neurological pathways in PD pathophysiology (Figure 3a).

To evaluate the validity of our network-based drug prediction approach, we first assessed its ability to recover known pharmacological treatments for PD. Several established PD medications exhibited negative proximity Z-scores across key progression endotype modules, supporting the biological relevance of our framework. For example, Levodopa showed consistently negative scores (motor: –1.15; MoCA: –1.56). Selegiline, a monoamine oxidase-B inhibitor used for PD and depression^101^, also demonstrated strong proximity (motor: –0.97; MoCA: –0.72). Similarly, Pramipexole, a dopamine agonist for motor symptom management^102^, exhibited negative Z-scores (motor: –0.50; MoCA: – 0.23). These findings provide a proof-of-concept that our proximity-based framework can recapture clinically effective PD drugs, thereby validating its potential to identify novel therapeutic candidates.

To further validate these candidates, we examined their associations with clinical outcomes using two large-scale, de-identified real-world patient datasets (RWD), focusing on PD-related complications (Table 2, Table S11). The INSIGHT Clinical Research Network includes longitudinal clinical records for approximately 21.6 million individuals from New York City and Houston. The OneFlorida+ Clinical Research Network encompasses data on nearly 17 million individuals in Florida, 2 million in Georgia, and 1 million in Alabama. A subset of 16 drugs was selected for analysis based on expert curation and the integration of multiple criteria (see Methods). We applied a computational trial emulation framework to estimate treatment effects from observational data. For each candidate drug, we defined the treated group as individuals who initiated the drug following a diagnosis of PD. Control groups were assembled using 1:1 nearest-neighbor propensity score matching among patients exposed to alternative treatments. Propensity scores were derived from baseline demographic and clinical covariates, and inverse probability of treatment weighting was used to adjust for residual confounding. Treatment effects were estimated using Cox proportional hazards models. In line with previous work, treatment benefit was defined as a reduced hazard of developing PD related outcomes.

**Table 2.** Real-word patient data summary statistics.

|  | INSIGHT | OneFlorida+ |
| --- | --- | --- |
| Number of patients | 44704 | 81063 |
| Age on index date, years; mean ± SD | 72.63 (10.30) | 71.17 (13.93) |
| Sex, n (%) |  |  |
| Female | 18565 (41.53) | 37559 (46.33%) |
| Male | 26139 (58.47) | 43501 (53.66%) |
| Unknown | 0 (0%) | 3 (0.00%) |
| Race, n (%) |  |  |
| White | 22834 (51.08) | 54206 (66.87%) |
| Black | 2790 (6.24) | 7617 (9.40%) |
| Asian | 2040 (4.56) | 869 (1.07%) |
| Other | 15756 (35.25) | 13440 (16.58%) |
| Unknown | 1284 (2.87) | 4931 (6.08%) |
| Ethnicity, n (%) |  |  |
| Hispanic | 4006 (8.96) | 14749 (18.20%) |
| Non-Hispanic | 27308 (61.09) | 55639 (68.64%) |
| Unknown | 13390 (29.95) | 10671 (13.16%) |

Among the 25 prioritized candidates, Zolpidem, Alprazolam, Duloxetine, and Primidone stood out with consistently strong network-based proximity scores across motor and cognition modules, as well as real-world evidence of clinical benefit. All four drugs demonstrated negative Z-scores across multiple endotypes, indicating close network proximity to disease modules and potential therapeutic relevance. Zolpidem, a sedative-hypnotic prescribed for insomnia, has been reported to improve motor function in selected PD patients ; our results showed strong enrichment (motor: -0.65, MoCA: -2.53). Alprazolam, a benzodiazepine indicated for anxiety and panic disorders, has been associated with improved gait stability and reduced freezing of gait in PD ; it ranked highly in our network (motor: -0.51, MoCA: -2.41). Duloxetine, a serotonin-norepinephrine reuptake inhibitor used for anxiety and chronic pain, has been shown to reduce OFF-time and improve motor symptoms in PD ; our method prioritized it with negative Z-scores (motor: -0.76). Primidone, an antiepileptic agent traditionally used for tremor management, particularly essential tremor, has also been studied as adjunctive therapy in parkinsonian tremor . It showed strong network-based relevance (motor: -1.08, MoCA: -2.79).

In real-world patient data, these four candidates were further associated with reduced risks of PD-related complications, particularly falls and dementia (Figure 3b). In the OneFlorida+ cohort, Zolpidem use was associated with a 21% lower risk of falls (HR = 0.79, 95% CI [0.77, 0.80]) and 18% lower risk of dementia (HR = 0.82, [0.80, 0.84]); similar protective effects were observed in INSIGHT (fall HR = 0.81, dementia HR = 0.55; all P < 0.001). Alprazolam was associated with 11% and 28% reduced risk for falls (HR = 0.89, [0.88, 0.90]) and dementia (HR = 0.72, [0.70, 0.74]) in OneFlorida+ and again showed consistent benefit in INSIGHT (fall HR = 0.82, dementia HR = 0.88; P < 0.001). Duloxetine was linked to a 12% and 14% reduction in fall (HR = 0.88, [0.85, 0.90]) and dementia risk (HR = 0.86, [0.83, 0.89]) respectively in OneFlorida+, with consistent directionality observed in INSIGHT). Primidone showed a modest 8% reduction in fall risk (HR = 0.92, [0.90, 0.94]) and a 38% reduction in dementia risk (HR = 0.62, [0.60, 0.64]) in OneFlorida+, with similar effects in INSIGHT (fall HR = 0.74, dementia HR = 0.71; P < 0.001). The effects of these drugs across additional endotypes are presented in Figure S5 and Table S11. While the magnitude of benefit varied between cohorts, potentially reflecting demographic and clinical heterogeneity, the consistent directionality of effects reinforces their potential for disease-modifying impact. Taken together, these results highlight Zolpidem, Alprazolam, Duloxetine, and Primidone as promising candidates for repurposing in PD, with the potential to attenuate both motor and cognitive progression.

To investigate potential mechanisms of action, we examined the overlap between drug-target networks and progression-endotype modules (Figure 3c). Notably, several genes appeared both as module genes and as direct targets of the prioritized drugs. For example, in the case of Zolpidem, the genes *AR* (associated with motor and MoCA deficits, and CSF levels of α-synuclein, Aβ_42_, phosphorylated tau, and total tau) and *GABRG2* (motor, MoCA) were identified as both module genes and direct targets. Similarly, for Alprazolam, *GABRG2* and *CREBBP* (motor, MoCA, α-synuclein, Aβ_42_, phosphorylated tau, and total tau) served dual roles. Beyond direct overlaps, we also observed indirect connections wherein module genes interact with target genes via intermediary nodes. For instance, *MDM2* (motor, MoCA, α-synuclein, Aβ_42_, phosphorylated tau, and total tau), a module gene for Zolpidem, was not a direct target but was linked to the target gene *GABRA2*. Similarly, for Alprazolam, *MAPK1* (motor, MoCA, α-synuclein, Aβ_42_, phosphorylated tau, and total tau) was connected to the target *GABRA1*. These observations suggest that both direct and network-mediated mechanisms may underlie the therapeutic effects of the candidate drugs.

## Discussion

Although the heterogeneity of PD progression is well recognized, its underlying mechanisms remain still unclear. In this study, we demonstrated that PD progression-endotypes modules provide a complementary strategy for identifying therapeutic opportunities. Specifically, we identified progression-endotype modules associated with distinct trajectories of PD progression relative to clinical symptoms or fluid biomarkers. To support the biological relevance of these modules, we performed pathway enrichment analysis and literature-based validation of module-associated genes. These analyses informed the development of a network-based drug repurposing framework, in which we quantified the proximity between PD endotype modules and known drug targets. By integrating computational screening, literature, clinical trial evidence, and real-world data, we prioritized 4 top candidate drugs with potential therapeutic relevance to PD.

We evaluated the robustness of our progression-associated gene signatures by replicating our framework in the independent PDBP cohort. Despite notable differences between PDBP and PPMI in follow-up duration (2.85 years versus 8.72 years on average), population characteristics, and medication data format (binary indicators in PDBP compared to continuous measures in PPMI), the replicated modules showed substantial overlap in gene composition (Figure S1). Several key clinical assessments available in PPMI, including GDS, STAI, and CSF biomarkers, were not present in the retrieved PDBP dataset. In addition, missing data were considerable; for example, MoCA scores were missing in 44.74% of cases, and REM sleep behavior assessments were missing in 99.22% of cases, leading to the exclusion of REM-related analyses from replication. Despite these limitations, the consistency of identified progression-related gene modules supports the robustness of our framework to variations in cohort characteristics and data availability when similar selection criteria are used.

Quantitative evaluation of the identified modules was conducted using network-based metrics such as node closeness and edge density (Figure S4). All endotype modules exhibited significantly shorter node closeness compared to random permutations (adjusted P < 0.05), and four modules (motor, MoCA, GDS, and autonomic) showed significantly higher edge density. In contrast, modules associated with fluid biomarkers such as α-synuclein and Aβ_42_ did not consistently demonstrate elevated density, possibly reflecting the peripheral network positioning of these biomarker-associated genes. These results highlight the importance of integrating both topological properties and biological annotations when evaluating disease modules.

To complement these quantitative findings, we performed a qualitative assessment of module genes and enriched pathways. In addition to known GWAS loci, we identified literature-supported genes and pathways relevant to PD pathophysiology. Endotype-specific genes may point to mechanisms linked to distinct clinical features. For example, *MCCC1* (specific in the depression endotype) is linked to leucine metabolism, and studies suggest leucine supplementation mitigates depressive behaviors, implicating *MCCC1* in mood-related PD symptoms. Several genes were found to be shared across motor and other endotype modules, suggesting their involvement in common molecular mechanisms underlying diverse aspects of PD progression. For instance, *ATP13A2* is shared by the motor, cognition, and ESS endotype modules. *ATP13A2* encodes a lysosomal P-type ATPase primarily localized to early and late endosomes and lysosomes, and mutations in *ATP13A2* are associated with autosomal recessive early-onset parkinsonism and are known to disrupt lysosomal homeostasis, impair autophagic flux, and promote α-synuclein aggregation^103^. The presence of *ATP13A2* across multiple symptom domains highlights its central role in neuronal proteostasis and oxidative stress responses relevant to both dopaminergic and non-dopaminergic systems.

Among the drug candidates identified, zolpidem emerged as a top-ranking drug, with its module genes showing proximity to zolpidem targets. zolpidem demonstrated consistently negative network proximity scores across all nine PD progression modules in our analysis, with Z-scores ranging from –2.6 to –0.5. Several studies have reported symptomatic benefits of zolpidem in PD, including improvements in motor and non-motor symptoms as well as abnormal involuntary movements. For example, a 1997 case report described a 61-year-old woman with a 25-year history of PD who showed substantial improvement in akinesia and rigidity following zolpidem administration^104^. More recent studies have corroborated these findings, suggesting potential therapeutic effects of zolpidem in selected PD populations (NCT03621046)^105,106^. However, conflicting evidence exists. A 1997 study questioned its indication for PD and noted increased side effects at higher doses^104^, and a 2014 population-based study suggested a dose-dependent increase in PD incidence with zolpidem use^107^. Our data highlight the importance of phenotype and network-based analyses for the identification of disease-modifying drugs that otherwise would be dismissed due to inconsistent results.

Beyond the four leading drug candidates, several additional high-ranking compounds from our network-based screen demonstrated potential therapeutic relevance. Rutin, a plant-derived flavonoid with antioxidant and neuroprotective properties, has been shown to protect dopaminergic neurons in PD models^108,109^. It exhibited strong proximity scores (motor: –1.73, MoCA: –1.20); however, its clinical applicability remains uncertain due to limited exposure in real-world data (n = 2). Warfarin, an anticoagulant that has completed Phase I clinical trials for PD, as registered on ClinicalTrials.gov (NCT02169440, NCT02305030), also ranked highly in our screen (motor: –1.72, MoCA: –4.46). Dequalinium, an antimicrobial agent proposed as a molecular probe for α-synuclein toxicity^110^, demonstrated strong proximity to PD modules (motor: –2.01, MoCA: –1.34). While literature supports its potential benefit, real-world validation was limited by sparse data on PD patients with relevant symptom profiles. In addition to these literature-supported drugs, we identified several drugs with no prior association with PD, yet strong negative proximity scores across endotype modules. Stavudine, an antiretroviral medication used in the treatment of HIV, has been reported to reduce NLRP3 inflammasome activation and promote amyloid-β autophagy in preclinical studies^111^. While these effects have been studied in the context of Alzheimer’s disease, the underlying mechanisms, including neuroinflammation and impaired protein clearance, are also relevant to Parkinson’s disease. These findings raise the possibility that Stavudine may exert neuroprotective effects in PD and merit further investigation. Axitinib, a tyrosine kinase inhibitor with anti-angiogenic and immunomodulatory properties, has not been evaluated in PD. However, dysregulation of immune signaling and vascular dysfunction have been implicated in PD progression, suggesting that Axitinib may also warrant exploration as a potential therapeutic candidate.

Mechanistically, the therapeutic effects of the top-ranking candidate drugs may be partly explained by their convergence on progression-relevant genes identified through our network framework. Several drugs, including directly target genes that also emerged as key module components, suggesting potential dual relevance in disease modulation. For instance, *AR* and *GABRG2*, targeted by Zolpidem and Alprazolam respectively, were shared across modules for motor, cognition, and associated with CSF biomarkers including α-synuclein and tau. In addition, drugs like Primidone and Duloxetine were found to target genes involved in neurotransmission, inflammation, or neurotrophic signaling, including *CHRNA7, HTR6, and AR*, further supporting their disease-modifying potential. Notably, we also observed indirect interactions between module genes and drug targets, suggesting that broader network-mediated effects may contribute to therapeutic efficacy. These findings highlight that network-based drug repurposing can capture not only proximity-based candidates but also potential mechanistic links between drugs and PD pathophysiology.

Our study offers several key strengths. First, we systematically investigated the molecular underpinnings of PD progression by constructing progression endotype modules that integrate clinical assessments, CSF biomarkers, whole-genome sequencing, and peripheral blood transcriptomic profiles. These modules not only reflect distinct biological drivers of PD progression but also support the hypothesis that different progression patterns are underpinned by distinct molecular mechanisms. The resulting modules were validated through both network topology and prior literature, reinforcing their biological plausibility. Importantly, module genes showed functional coherence at the pathway level, and several were enriched in known PD-relevant biological processes, providing further biological validation. Second, by incorporating high-confidence drug–target interactions from curated databases, we applied a network-based proximity framework to identify repurposable drugs with potential to modify PD progression. Among the top-ranked candidates, some were supported by prior studies or clinical trials, while others represent novel findings that may warrant further investigation. Finally, we evaluated candidate drugs using real-world clinical data from two large independent cohorts, demonstrating associations with reduced risks of cognitive and motor decline, thereby highlighting the translational relevance and robustness of our integrative framework.

Despite its strengths, our study has several limitations. First, the network proximity–based drug repurposing approach is constrained by the completeness and accuracy of existing drug–target interaction databases, which remain inherently incomplete and may fail to fully capture the therapeutic landscape. Second, rather than conducting a genome-wide screen, our genetic analysis focused on previously reported PD-associated variants from recent GWAS, potentially overlooking additional relevant loci that contribute to specific PD progression modules. Given the greater number of known AD loci compared to PD, this imbalance may also inflate the apparent overlap between AD and PD modules. Third, although transcriptomic data were integrated, the lack of additional tissue types and high-resolution modalities such as brain sample and single cell sequencing data limits the mechanistic granularity of our findings. Moreover, the use of real-world data introduces inherent noise, including potential diagnostic inaccuracies. Finally, while current data quality and availability may limit the resolution of fully distinct molecular signatures across all progression endotype modules, the reproducibility of shared features across independent cohorts supports the overall robustness of our framework. Future integration of higher-resolution molecular data may help delineate endotype-specific signals more precisely.

In summary, this study presents a novel integrative approach to define PD progression endotypes, characterize their molecular architecture, and enable drug repurposing through a network-based framework. The identification of candidate drugs with supporting evidence from literature and real-world data illustrates the potential of this strategy to guide future translational efforts for Parkinson’s disease and establishes a framework that could be applied to other complex brain disorders.

## Data Availability

All data produced in the present study are available from the PPMI study - https://www.ppmi-info.org/access-data-specimens/download-data

